# Mathematical Modeling of Rift Valley Fever in the Sahelian Zone

**DOI:** 10.64898/2026.07.15.26358164

**Authors:** H. Djimramadji, Bienvenu Ndonane, P. Djaouga, H. Markhous Mahamat, E. Djoumountanan, K. Tobaye, F. Mahamat Abakar

## Abstract

We develop a mathematical model of Rift Valley Fever integrating mosquito vectors, ruminants, and humans, based on an *SEIR*-type structure with vertical transmission in vectors. Local data from the Sudanian and especially the Sahelian zones are used to capture the impact of climatic variations on mosquito population dynamics. The mathematical analysis establishes the model’s positivity, determines the basic reproduction number *R*_0_, and demonstrates the local and global stability of the disease-free equilibrium. Sensitivity analysis (PRCC) highlights the most influential parameters, while the stochastic approach using a continuous-time Markov chain confirms the major role of seasonal rainfall. Numerical simulations reveal a peak in animal and human infections around the 9^th^ month, correlating with periods of heavy rainfall. This model provides a relevant tool for surveillance and prevention within a “One Health” approach in Chad.

## 1 Introduction

Rift Valley Fever (RVF) is a major vector-borne zoonosis, an infectious, virulent, and inoculable disease caused by a virus of the genus *Phlebovirus* in the family *Bunyaviridae* [11].

The RVF virus was first isolated in sheep in the Rift Valley of Kenya in 1930 [11]. RVF affects both domestic and wild ruminants.

The virus is transmitted to animals by mosquito vectors belonging to the genera *Culex, Aedes, Anopheles, Eretmapodites*, and *Mansonia* [11]. Animals remain infectious during the viremic period, which lasts from 6 to 18 hours, and can extend up to six to eight days [11].

Transmission can also occur from mosquito to mosquito through eggs [11]. Female mosquitoes pass the virus to their offspring via eggs, in which the virus remains infectious for several years. Eggs hatch during heavy rainfall, leading to a rapid increase in mosquito populations and accelerated virus transmission to animals and humans.

In humans, infection can occur through:

- mosquito bites;
- bites from blood-feeding flies [17];
- contact with blood, bodily fluids, or tissues of infected animals, notably during slaughtering or animal care [4].

In animals, RVF manifests in three forms [18]:

- Hyperacute: severe form with anorexia, lethargy, depression; lambs are unable to stand; 94 % mortality within 24 hours.
- Acute: anorexia, hyperthermia, high fever, bloody diarrhea in young animals; 90 % mortality within two to three days. Frequent abortions accompanied by metritis and perimetritis. Some females die without symptoms (mortality up to 20 %).
- Subacute: observed in both young and adult animals. Common symptoms include abortion, anorexia, fever, decreased lactation, bristled coat, excessive salivation, and foul diarrhea.

Common signs: mucopurulent nasal discharge, depression, muscle spasms, locomotor disorders, jaundice.

In humans, the disease is often mild. After an incubation period of 3 to 6 days, it can be asymptomatic or present with [18]:

- acute fever,
- muscle pain,
- fatigue, anorexia, headache,
- nausea, photosensitivity,
- hemorrhagic complications, neurological signs, vision loss.

Death can occur in severe cases.

Prophylactic measures exist, and some treatment options are being explored; however, there is no specific vaccine or antiviral therapy for RVF. Treatment is symptomatic.

Chad, due to its ecological and pastoral conditions, is a high-risk area for virus circulation. Domestic species affected by RVF in Chad include cattle, sheep, goats, and camelids. Humans are also at risk. Constant proximity between herders and animals, along with pastoral nomadism, increases the risk of transmission, particularly when herds move to high-risk areas where mosquitoes can also migrate.

The first serological evidence of RVF virus in ruminants in Chad dates back to 1967. A serological study on sheep and wild ruminants revealed longstanding viral circulation in the region [12].

In 2002, a serological survey conducted during the rainy season indicated active virus transmission among domestic ruminants, confirming the presence of antibodies in many animals [19].

More recently, a 2014 seroprevalence study along the shores of Lake Chad found that 37.8 % of cattle, 18.8 % of goats, and 10.8 % of sheep had anti-RVF antibodies, confirming high viral circulation in this region [3].

In 2019, nine cases were confirmed in gazelles (*Oryx gazella*) in the Wadi Rimé Reserve in Batha, eight of which were fatal, highlighting active virus transmission in wildlife [1].

Human exposure to RVF virus has also been documented. In 2003, French military personnel deployed in Chad were infected, providing evidence of active human transmission in certain regions [6].

A more recent study conducted from 2018 to 2023 in the Yao and Danamadji districts investigated the sero-prevalence of RVF, brucellosis, and Q fever in humans and animals. Results highlighted several risk factors, including close contact with animals, lack of proper sanitary practices, and transhumance movements [2].

Mathematical models of RVF have been proposed in several African contexts. Gaff et al. [8] developed a SEIR/SEI model incorporating vertical transmission in mosquitoes and the effect of rainfall on egg hatching. Xue et al. [22] extended this approach with a multi-host stochastic model. However, these models often focus on East African contexts (Kenya, Tanzania), without accounting for the specificities of Sahelian nomadic livestock systems. Chuma [5] implemented seasonal rainfall effects in transmission rates, confirming their importance in epidemic cycles in tropical humid climates. Oguntolu et al. [16] focused on human-livestock interactions, using an ODE model applied to African contexts centered on cattle farming. Mpeshe [14] applied fractional derivatives to model peri-domestic urban dynamics, adding complexity to the analysis of the infected population and *R*_0_.

These contributions considerably enrich the methodological toolkit. For the Chadian context, our approach incorporates:

- pastoral transhumance as an epidemiological factor,
- dynamics specific to Chad’s climatic zones (Sahelian and Sudanian regions),
- and explicit interactions among animals, humans, vectors, climate, and livestock practices.

Additionally, our model differs from previous approaches through several innovative features:

- It accounts for the effect of seasonal rainfall on vector mosquito emergence via delayed hatching of infected eggs, a crucial factor in epidemic initiation.
- It explicitly incorporates direct virus transmission to humans through contact with infected animals, reflecting local slaughtering, care, and consumption practices.
- It adopts an integrated One Health approach, linking animal, human, vector, and climatic dynamics within a single mathematical framework.

Consequently, this model aims to provide a tool adapted to the epidemiological and pastoral realities of Chad, supporting prevention, surveillance, and rapid response strategies against RVF in vulnerable areas. It seeks to fill a methodological gap in modeling vector-borne zoonoses in Central Africa under a One Health framework.

## 2 Mathematical Modeling of Rift Valley Fever

### 2.1 Biological Assumptions

Rift Valley Fever (RVF) is a viral zoonosis primarily transmitted by infected mosquitoes (species *Aedes, Culex, Anopheles*, etc.) to animals (cattle, sheep, goats, camelids) and humans. Transmission can also occur through:

- mosquito-to-mosquito vertical transmission (infected eggs),
- mosquito-to-human or mosquito-to-animal via bites,
- direct animal-to-human contact with blood or contaminated animal products.

Rainfall triggers hatching of infected eggs and mosquito population proliferation, influencing the epidemic dynamics.

### 2.2 Mathematical Formulation of the Model

The proposed model describes the transmission dynamics of Rift Valley Fever (RVF) among three populations: mosquitoes (vectors), animals (ruminants), and humans. It is a compartmental SEIR-type system adapted for each group, incorporating vertical transmission in mosquitoes and the effect of rainfall on vector demography.

### 2.3 Model Description

The proposed model is a compartmental *SEIR* system adapted to three interacting populations: mosquito vectors, animals (ruminants), and humans. Each compartment represents an epidemiological state (Susceptible *S*, Exposed *E*, Infectious *I*, Recovered *R*), with an additional compartment for infected eggs (*O*) in mosquitoes to account for vertical transmission.

- Vectors (mosquitoes): Vector dynamics are represented by the classes *S*_v_, *E*_v_, *I*_v_, and *O*. Mosquitoes are born at a rate Λ_v_(*t*) dependent on seasonal rainfall, can become infected upon contact with infected animals, and transmit the virus to ruminants and humans. A proportion *θ* of eggs laid by infected mosquitoes are contaminated, ensuring virus persistence during inter-epidemic periods.
- Animals (ruminants): Animals are divided into *S*_a_, *E*_a_, *I*_a_, and *R*_a_. They become infected following bites from infected mosquitoes and can develop acute, subacute, or hyperacute forms. The *I*_a_ compartment accounts for disease-induced mortality (*d*_a_) in addition to natural mortality. Recovered animals acquire long-lasting immunity.
- Humans: Humans (*S*_h_, *E*_h_, *I*_h_, *R*_h_) can be infected either by bites from infected mosquitoes (*I*_v_) or through direct contact with infected animals (*I*_a_), particularly during livestock handling, slaughtering, or care activities. Humans do not exhibit vertical transmission.

The compartmental structure is illustrated in Figure 1.

**Figure 1:**
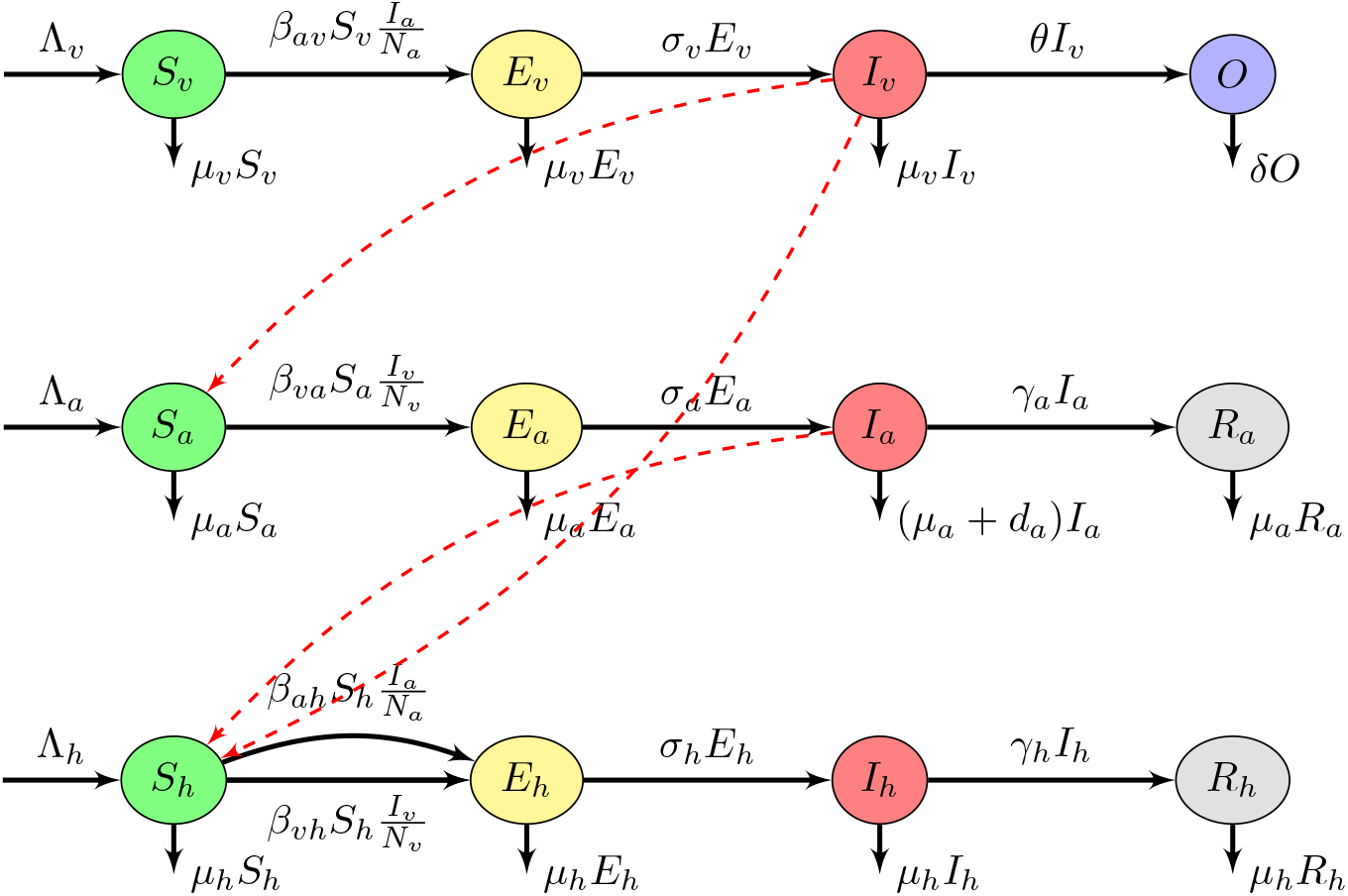
Flow diagram of the vector-borne model illustrating transmission among mosquitoes, ruminants, and humans.

#### 2.3.1 Effect of Rainfall

Vector dynamics are influenced by rainfall *P* (*t*), which affects the mosquito birth rate. The hatching of infected eggs can be modeled by a rainfall-dependent function:

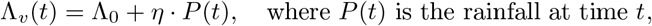

where Λ_0_ is the baseline rate and *η* is a sensitivity parameter (Sensitivity coefficient of mosquito birth rate to rainfall).

To model a more realistic behavior, especially during heavy rainfall that may destroy larval habitats (flooding), we use a saturating function inspired by Michaelis-Menten or Holling II type models, commonly used in ecology to represent limiting effects [7]:

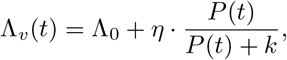

where:

- *k* is a threshold parameter controlling the saturation point (rainfall half-saturation constant).

This function is increasing but plateaus when *P* (*t*) ≫ *k*, reflecting a biologically limited response. The resulting mosquito birth rate as a function of rainfall is shown in Figure 2.

**Figure 2:**
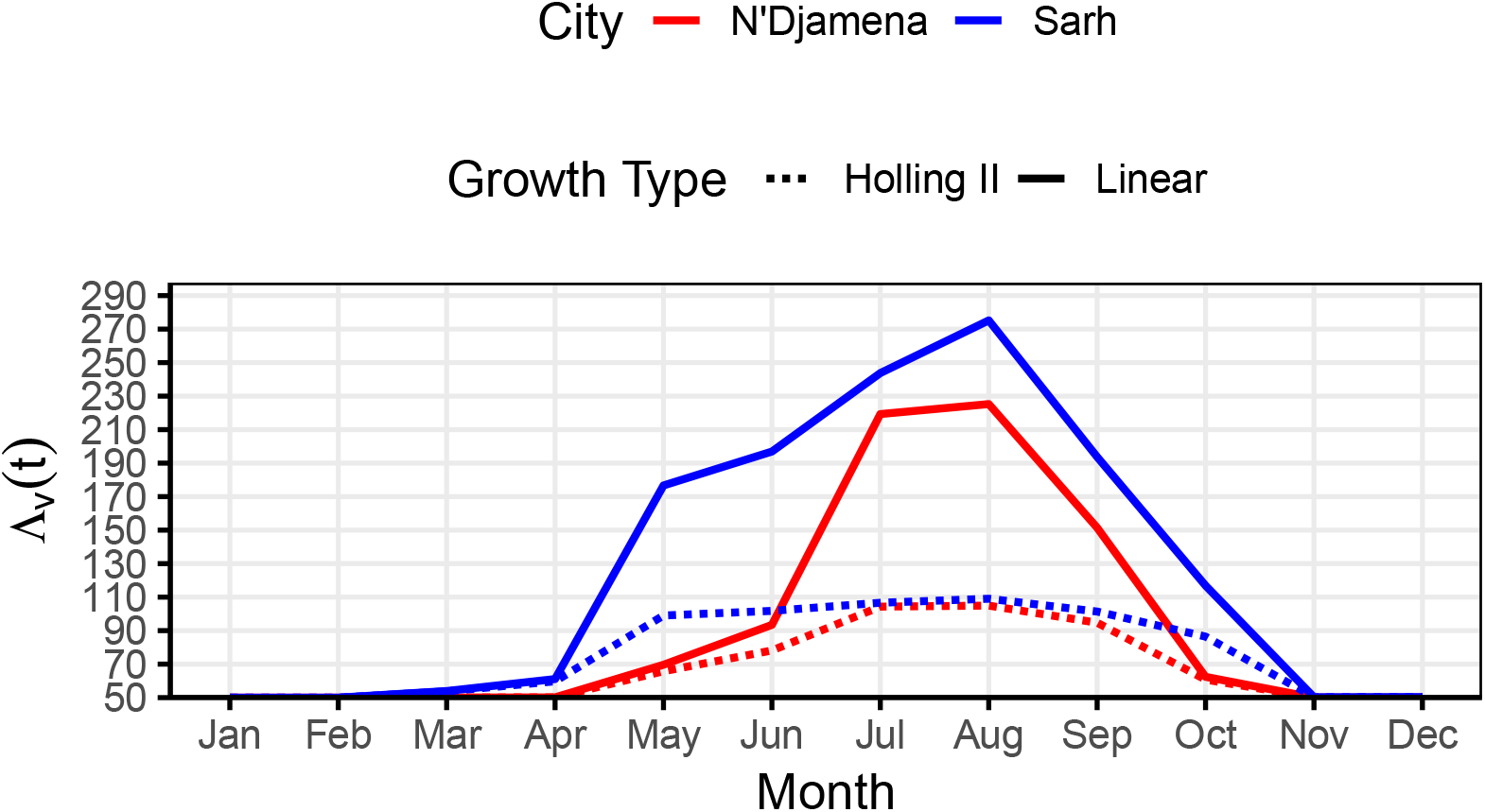
Comparison of mosquito birth growth in the cities of N’Djamena and Sarh from 2010 to 2020 as a function of rainfall. Source of rainfall data: National Meteorology Agency (ANAM-TCHAD, 2021).

#### 2.3.2 Mosquito Population

Mosquitoes are divided into three compartments: Susceptible (*S*_v_), Exposed (*E*_v_), and Infectious (*I*_v_). An additional compartment *O* represents infected eggs laid by infected mosquitoes. Mosquitoes become infected upon contact with infected animals. Infected eggs are laid at a proportion *θ* and have their own mortality rate *δ*.

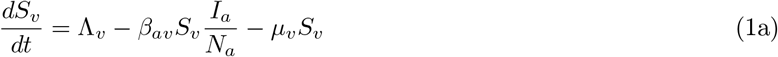

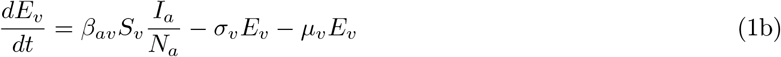

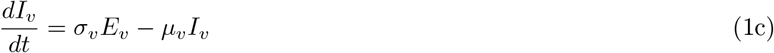

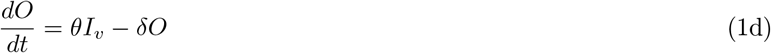

#### 2.3.3 Animal Population (Ruminants)

Animals also follow an SEIR structure, with infection via bites from infected mosquitoes. Vertical transmission is not considered in animals, but disease-specific mortality is included through the parameter *d*_a_.

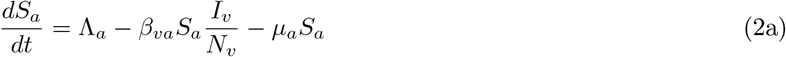

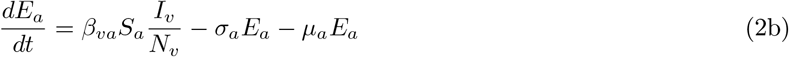

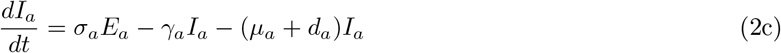

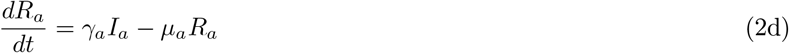

#### 2.3.4 Human Population

Humans can become infected either through bites from infected mosquitoes or by direct contact with infected animals. The compartmental structure is also of the SEIR type.

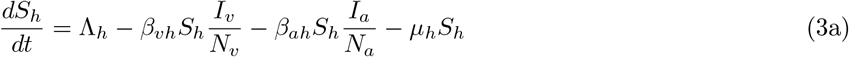

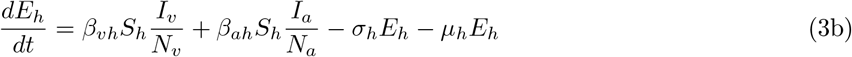

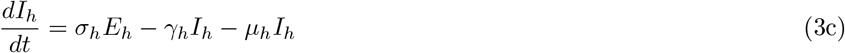

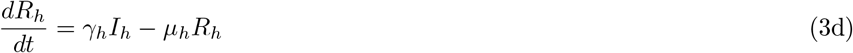

The complete set of parameters is summarized in Table 1.

**Table 1:** Model parameters and their biological definitions.

| Parameter | Description |
| --- | --- |
| $\Lambda_v$ | Mosquito birth rate |
| $k$ | Rainfall half-saturation constant |
| $\eta$ | Sensitivity coefficient of mosquito birth rate to rainfall |
| $\beta_{av}$ | Transmission rate from animal to vector |
| $\beta_{va}$ | Transmission rate from vector to animal |
| $\beta_{vh}$ | Transmission rate from vector to human |
| $\beta_{ah}$ | Transmission rate from animal to human (direct contact) |
| $\mu_v, \mu_a, \mu_h$ | Mortality rates of mosquitoes, animals, humans |
| $\sigma_v, \sigma_a, \sigma_h$ | Progression rates to infectiousness |
| $\gamma_a, \gamma_h$ | Recovery rates |
| $\theta$ | Proportion of infected eggs laid by infected mosquitoes |
| $\delta$ | Mortality rate of eggs |
| $d_a$ | Disease-induced mortality rate in animals |
| $N_v, N_a$ | Total populations of vectors and animals |

### 2.4 Combined System

In this section, we present the general model resulting from the combination of the previous models.

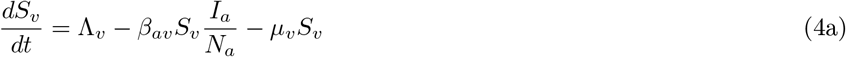

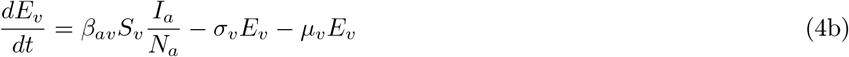

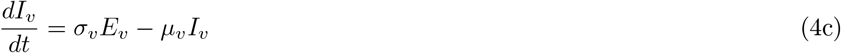

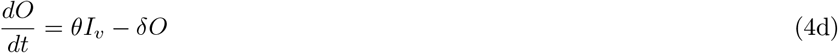

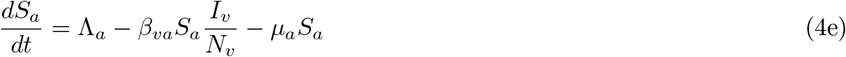

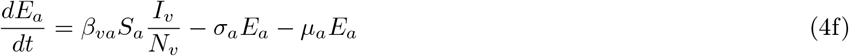

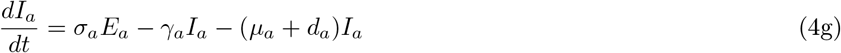

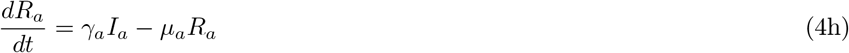

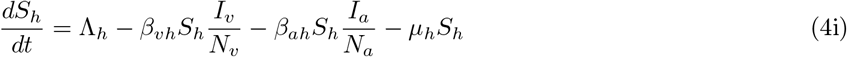

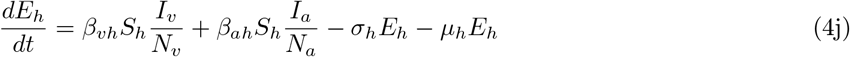

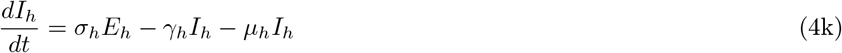

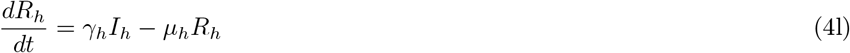

subject to the initial conditions:

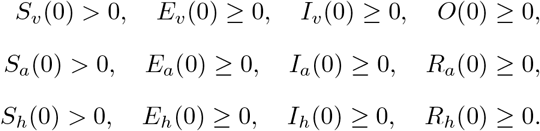

## 3 Mathematical Analysis of the Model

### 3.1 Existence and Uniqueness of the Solution

Consider the full system combining the mosquito, animal, and human populations in vector form:

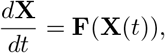

where

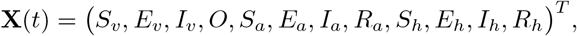

with positive initial conditions

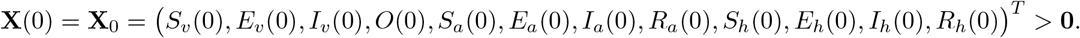

The function **F** : ℝ ^12^ → ℝ ^12^ is defined by the differential system given in equations (4), which is composed of linear combinations, products, and quotients where the denominators *N*_v_ = *S*_v_ + *E*_v_ + *I*_v_ and *N*_a_ = *S*_a_ + *E*_a_ + *I*_a_ + *R*_a_ remain strictly positive in a neighborhood of the initial condition.

Since **F** is locally Lipschitz continuous (its components are differentiable and the fraction terms are well-defined for *N*_v_, *N*_a_ *>* 0), the Cauchy-Lipschitz theorem [13] ensures the existence and uniqueness of a solution **X**(*t*) defined locally on an interval [0, *T*_max_).

The system is an epidemiological model with positive birth, transmission, recovery, and mortality rates. By construction, solutions originating from strictly positive initial conditions remain in 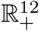 for all *t* ∈ [0, *T*_max_).

Therefore, for any initial condition **X**_0_ *>* **0**, there exists a unique local positive solution of the system modeling the dynamics of mosquito, animal, and human populations.

#### Lemma 3.1.

*The solution* **X**(*t*) *of system* (4) *remains strictly positive for all t* ≥ 0.

*Proof*. We rewrite equations (4a) (4e), and (4i) as

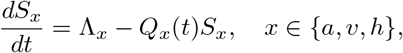

where 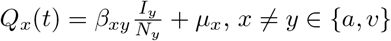, and 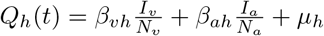.

The solution is

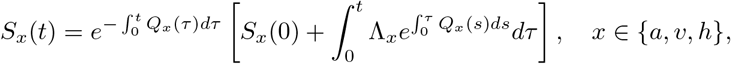

so that *S*_x_(*t*) ≥ 0 for all *t* ≥ 0.

Similarly, for equations (4b) and (4f), we have

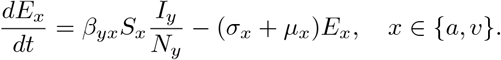

By contradiction, suppose there exists *t*_1_ *>* 0 such that *E*_x_(*t*_1_) = 0 and *E*_x_(*t*) *>* 0 for all *t* ∈ [0, *t*_1_) while all other components of **X**(*t*_1_) *>* 0. Then, evaluating the derivative at *t*_1_ gives

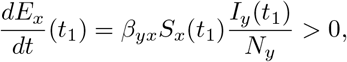

which implies that *E*_x_(*t*) cannot drop below zero. Thus, *E*_x_(*t*) ≥ 0 for all *t* ≥ 0.

The same argument applies for *E*_h_(*t*) in (4j) and for the other compartments:

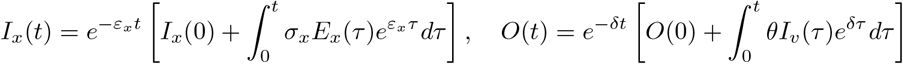

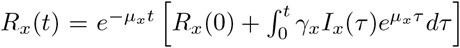, with 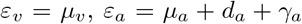, and 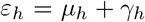,. Hence, all components of **X**(*t*) remain strictly positive (or non-negative if initially zero) for all *t* in the existence interval of the solution.

#### Lemma 3.2.

*The solutions of system* (4) *remain bounded for all t* ≥ 0 *in the positively invariant set*

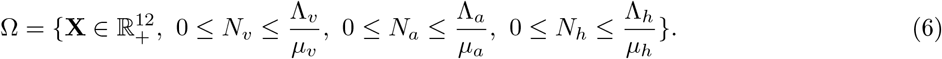

*Proof*. To show that the solutions remain bounded for all *t* ≥ 0, consider the total population in each group:

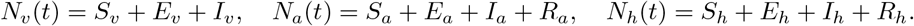

Summing the corresponding equations yields:

For mosquitoes:

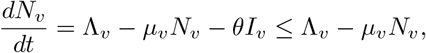

which has the explicit solution

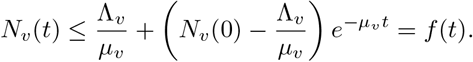

- If 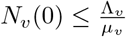, then 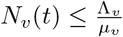 for all *t >* 0.
- If 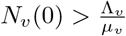, then *f* (*t*) is decreasing and *N*_v_(*t*) *< f* (*t*) *< f* (0) = *N*_v_(0) for all *t >* 0.

Thus, 0 ≤ *N*_v_(*t*) ≤ max(*N*_v_(0), Λ_v_*/µ*_v_), so *N*_v_(*t*) is bounded. Similarly,

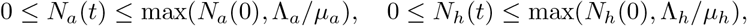

so both *N*_a_(*t*) and *N*_h_(*t*) are bounded.

Hence, the total populations of vectors, animals, and humans remain bounded for all *t* ≥ 0, ensuring that the solutions stay in the physically meaningful and bounded domain Ω defined in (6).

### 3.2 Disease-Free Equilibrium

The disease-free equilibrium corresponds to the state where there are no exposed or infected individuals in any population. In other words:

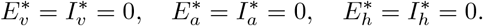

At equilibrium, the system reduces to:

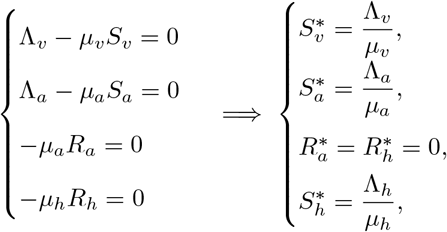

Thus, the disease-free equilibrium is given by

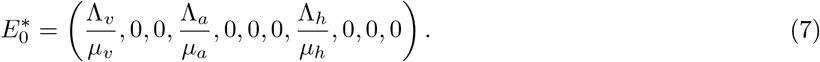

This point represents the healthy state where no infection is present in the populations.

### 3.3 Basic Reproduction Number

To study the stability of the disease-free equilibrium, we calculate the basic reproduction number R_0_ using the next-generation matrix method by Van den Driessche and Watmough [21].

Define the vector of infectious variables

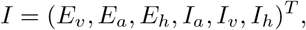

and construct the vectors F and −V representing, respectively, the new infections entering *I* and the transitions within and out of *I*:

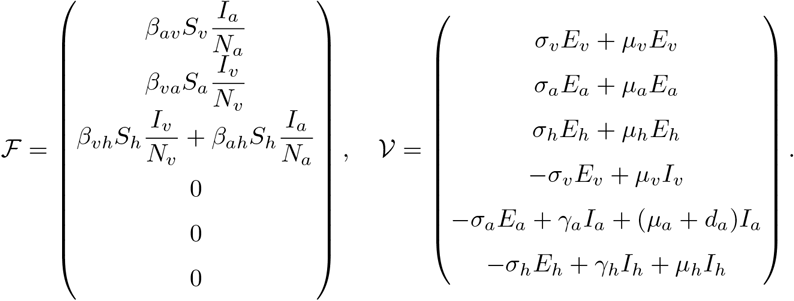

At the disease-free equilibrium 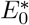 (7), the Jacobian matrices of new infections *F* and transitions *V*, defined by

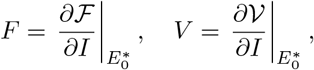

are given by:

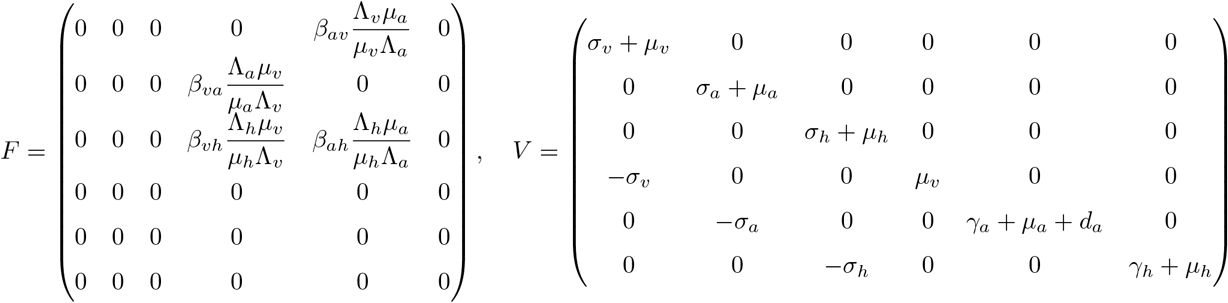

The basic reproduction number R_0_ is the spectral radius (dominant eigenvalue) of the next-generation matrix:

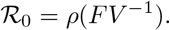

Since *V* is block-diagonal with strictly positive diagonal entries, it is invertible. Its inverse is obtained by inverting the 2×2 blocks:

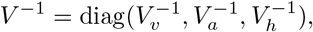

with

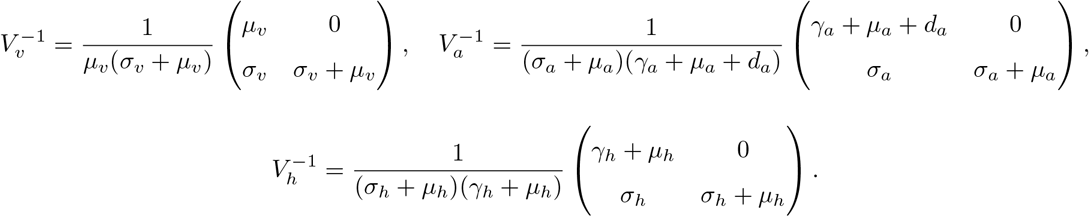

Hence,

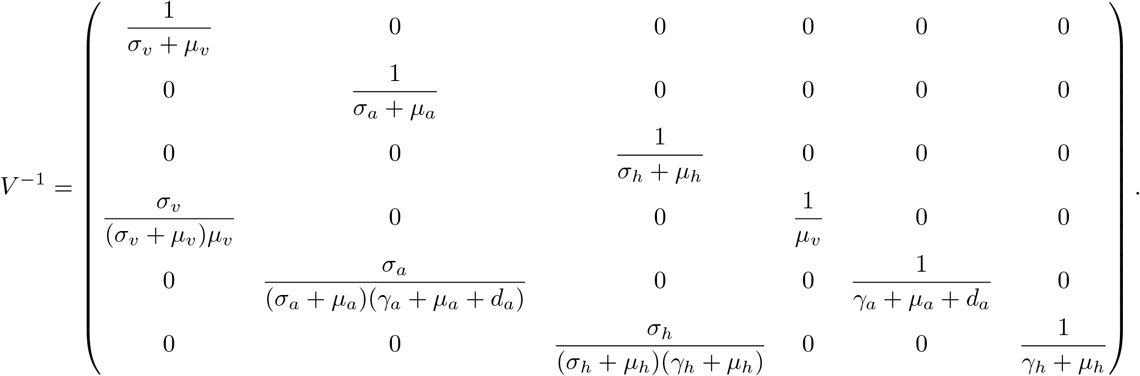

The next-generation matrix is then:

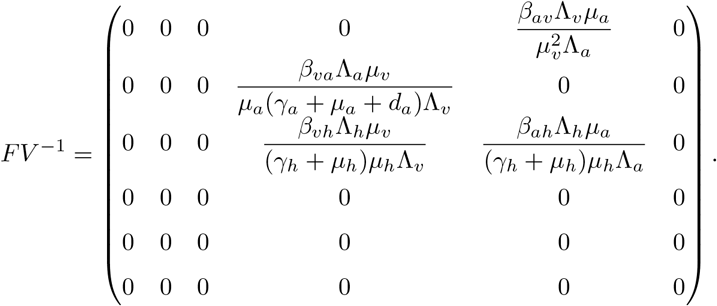

The basic reproduction number *R*_0_ is given by the spectral radius of *FV* ^−1^, that is:

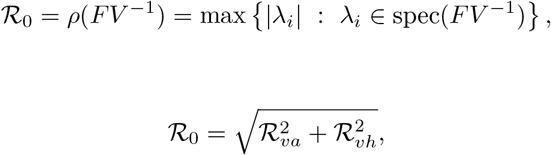

where

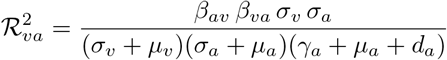

and

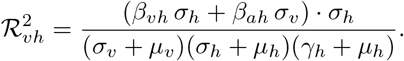

### 3.4 Stability of the Disease-Free Equilibrium

At the disease-free equilibrium, i.e., when *I*_a_ = *I*_h_ = 0, we have:

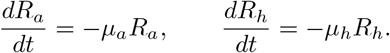

The associated solutions are exponentially decreasing:

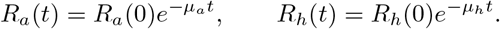

Thus,

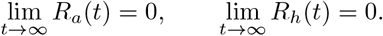

#### Theorem 3.3.

*The disease-free equilibrium* 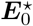 *of* (7) *is locally asymptotically stable if* R_0_ *<* 1 *and unstable if* R_0_ *>* 1.

*Proof*. By construction of the matrices *F* and *V*, assumptions A1–A4 of Theorem 2 in [21] are satisfied.

To verify assumption A5, consider system (4) in the absence of disease, i.e., setting all infectious variables to zero:

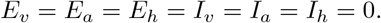

The system reduces to:

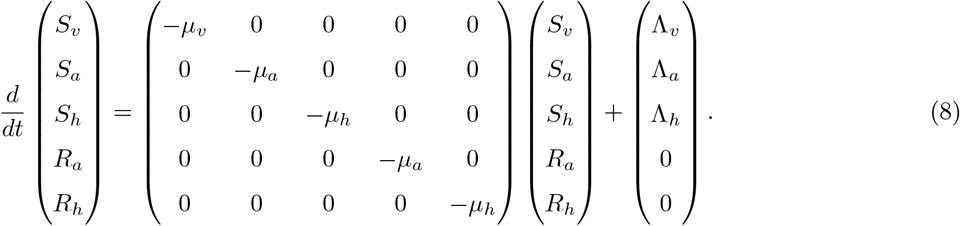

Let

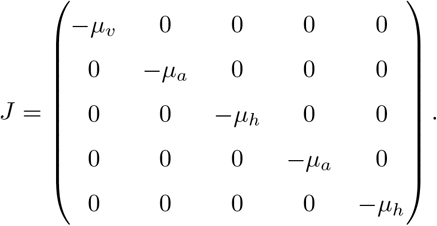

Matrix *J* is diagonal with negative entries, hence invertible with strictly negative eigenvalues. It thus admits a unique equilibrium 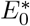 (7), which is globally asymptotically stable since

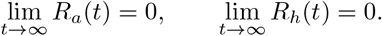

Therefore, assumption A5 of Theorem 2 in [21] is satisfied, and we can conclude that R_0_ *<* 1 determines the stability of the disease-free equilibrium (7).

### 3.5 Global Stability of the Disease-Free Equilibrium

#### Theorem 3.4.

*The disease-free equilibrium* 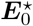 *of* (7) *is globally asymptotically stable if* R_0_ *<* 1. *Proof*. Consider the Lyapunov function [10]:

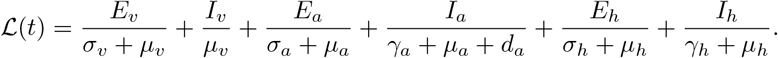

Differentiating L(*t*) along solutions of the system gives:

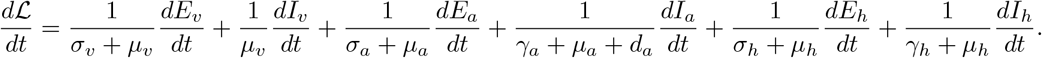

By substituting the system derivatives, we obtain

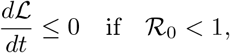

with equality only at the disease-free equilibrium (7). By LaSalle’s invariance principle [9], the disease-free equilibrium 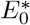 is globally asymptotically stable when R_0_ *<* 1.

## 4 Stochastic Analysis of the Model

In the numerical analysis, we use a continuous-time Markov chain (CTMC) analogue of the system to study infection events. The chain is defined based on the transitions and rates listed in Tables 2 and 3, representing mosquitoes and animals, and humans, respectively.

**Table 2:** Transitions and rates for the CTMC of animals and mosquitoes.

| Transition | Event $x, y \in \{a, v\}, x \neq y$ | Rate |
| --- | --- | --- |
| $S_x \rightarrow S_x + 1$ | Birth of a susceptible $x$ | $\Lambda_x$ |
| $S_x \rightarrow S_x - 1$ | Infection by an infectious $y$ | $\beta_{yx} S_x \frac{I_y}{N_y}$ |
| $S_x \rightarrow S_x - 1$ | Natural death of $S_x$ | $\mu_x S_x$ |
| $E_x \rightarrow E_x + 1$ | Infection (entering $E_x$) | $\beta_{yx} S_x \frac{I_y}{N_y}$ |
| $E_x \rightarrow E_x - 1$ | Progression to $I_x$ | $\sigma_x E_x$ |
| $E_x \rightarrow E_x - 1$ | Natural death of $E_x$ | $\mu_x E_x$ |
| $I_x \rightarrow I_x + 1$ | Progression from $E_x$ | $\sigma_x E_x$ |
| $I_a \rightarrow I_a - 1$ | Recovery of $I_a$ | $\gamma_a I_a$ |
| $I_a \rightarrow I_a - 1$ | Disease-induced death | $d_a I_a$ |
| $I_x \rightarrow I_x - 1$ | Natural death of $I_x$ | $\mu_x I_x$ |
| $R_a \rightarrow R_a + 1$ | Recovery from $I_a$ | $\gamma_a I_a$ |
| $R_a \rightarrow R_a - 1$ | Natural death of $R_a$ | $\mu_a R_a$ |

**Table 3:** Transitions and rates for the CTMC of humans.

| Transition | Event | Rate |
| --- | --- | --- |
| $S_h \rightarrow S_h + 1$ | Birth of a susceptible human | $\Lambda_h$ |
| $S_h \rightarrow S_h - 1$ | Infection by $I_v$ or $I_a$ | $\left( \beta_{vh} \frac{I_v}{N_v} + \beta_{ah} \frac{I_a}{N_a} \right) S_h$ |
| $S_h \rightarrow S_h - 1$ | Natural death of $S_h$ | $\mu_h S_h$ |
| $E_h \rightarrow E_h + 1$ | Infection (entering $E_h$) | same as above |
| $E_h \rightarrow E_h - 1$ | Progression to $I_h$ | $\sigma_h E_h$ |
| $E_h \rightarrow E_h - 1$ | Natural death of $E_h$ | $\mu_h E_h$ |
| $I_h \rightarrow I_h + 1$ | Progression from $E_h$ | $\sigma_h E_h$ |
| $I_h \rightarrow I_h - 1$ | Recovery of $I_h$ | $\gamma_h I_h$ |
| $I_h \rightarrow I_h - 1$ | Natural death of $I_h$ | $\mu_h I_h$ |
| $R_h \rightarrow R_h + 1$ | Recovery from $I_h$ | $\gamma_h I_h$ |
| $R_h \rightarrow R_h - 1$ | Natural death of $R_h$ | $\mu_h R_h$ |

## 5 Numerical Simulations

In this section, we perform a sensitivity analysis of R_0_ (see Section 5.2) and deterministic simulations (see Section 5.4) to illustrate our mathematical analysis.

### 5.1 Calibration Method

Parameter calibration was performed using maximum likelihood estimation combined with a continuous-time Markov chain (CTMC) approach. The objective function minimized the weighted sum of squared errors between model predictions and field data:

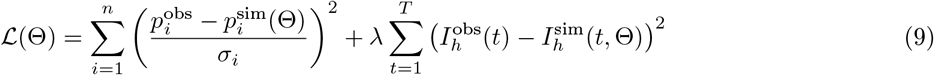

where 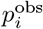 are observed seroprevalence data from [3] (cattle: 37.8%, goats: 18.8%, sheep: 10.8%), 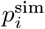 are model-predicted prevalences at equilibrium, *σ*_i_ are standard errors, and *λ* is a weighting factor for human incidence data from [2].

Optimization was performed using the optimx package in R [15] with multiple algorithms (L-BFGS-B, nlminb, bobyqa) to ensure global convergence. The 95% confidence intervals were obtained via Markov Chain Monte Carlo (MCMC) using the FME package [20], with 5,000 iterations after a burn-in period of 1,000.

The rainfall-dependent mosquito birth rate was modeled as:

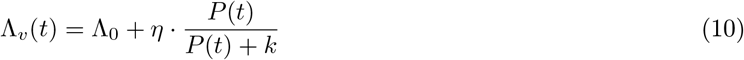

where *P* (*t*) is monthly rainfall from ANAM-TCHAD data (2010–2020) and *k* = 30 mm is the half-saturation constant. The calibrated values are reported in Table 4.

**Table 4:**
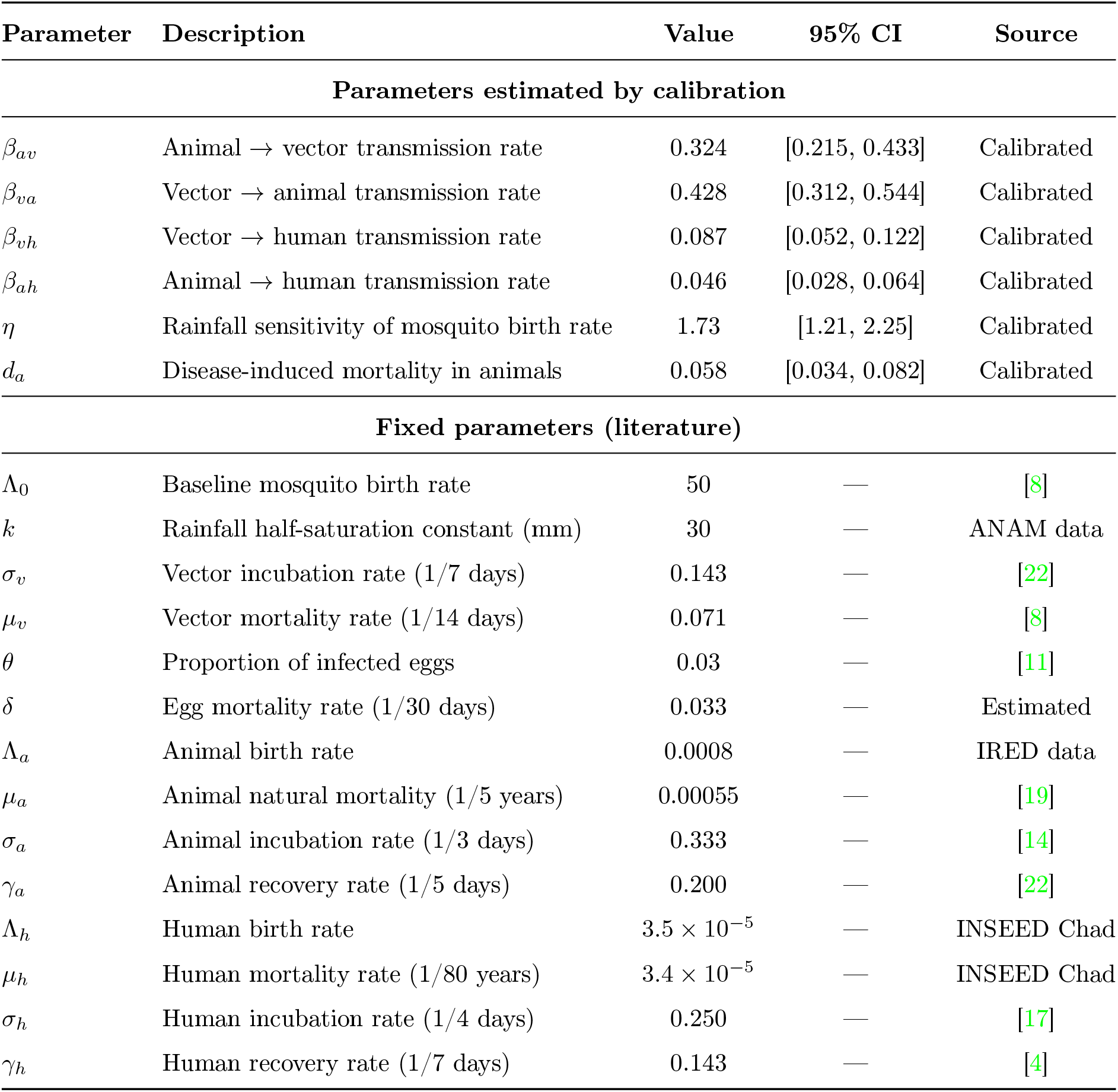
Calibrated parameter values for the Rift Valley Fever model in Chad.

### 5.2 PRCC Sensitivity Analysis

The sensitivity of the model was evaluated using the Partial Rank Correlation Coefficient (PRCC) to identify the parameters that most influence the dynamics of Rift Valley Fever. The results indicate that the number of infectious (*I*_v_) and exposed (*E*_v_) mosquitoes is strongly correlated with mosquito birth sensitivity to precipitation (*η*) and the transmission rate from animals to vectors (*β*_av_). For ruminants, the exposed (*E*_a_) and infectious (*I*_a_) compartments are mainly influenced by the vector-to-animal transmission rate (*β*_va_) and disease-induced mortality (*d*_a_). Human infections (*I*_h_) are sensitive to both vector-to-human (*β*_vh_) and animal-to-human (*β*_ah_) transmission rates. The Partial Rank Correlation Coefficient (PRCC) analysis is summarized in Figure 3.

**Figure 3:**
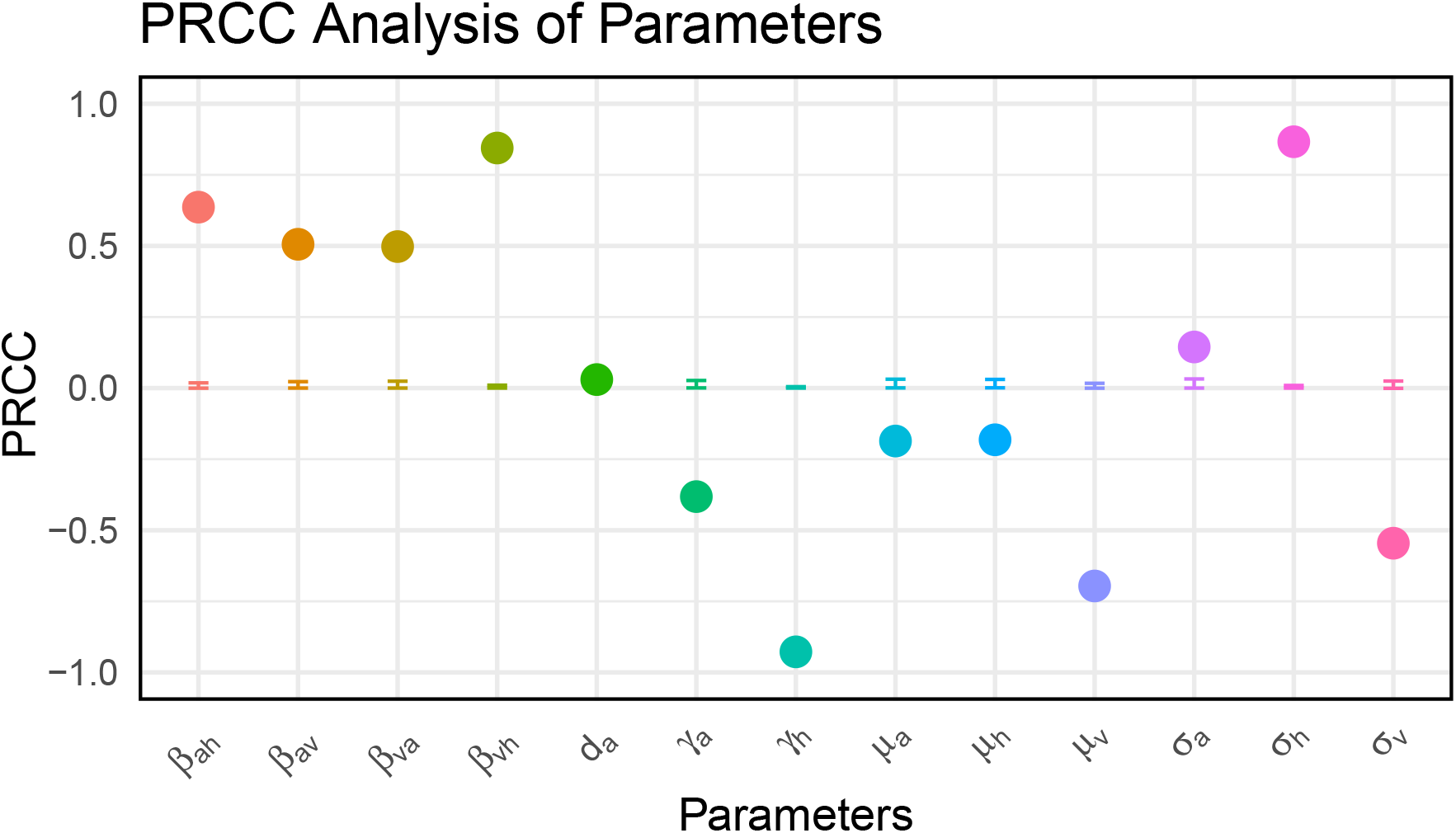
Sensitivity analysis of the basic reproduction number R_0_ using PRCC.

### 5.3 Stochastic Simulation

Stochastic simulations based on the CTMC illustrate the dynamics of Rift Valley Fever among animals and mosquitoes 2 in Chad, and humans 3. The CTMC simulation results are presented in Figures 4 and 5. The susceptible mosquito population starts high and varies slightly over time, reflecting both birth of new mosquitoes and transition to the infectious state after contact with infected animals (Figs. 4a and 5a). Infectious mosquitoes, initially few, increase rapidly to reach a peak, corresponding to the most intense period of virus transmission to animals (Figs. 5a and 5b). This critical phase is influenced by the rainy season and vector density, characteristic of the Chadian context.

**Figure 4:**
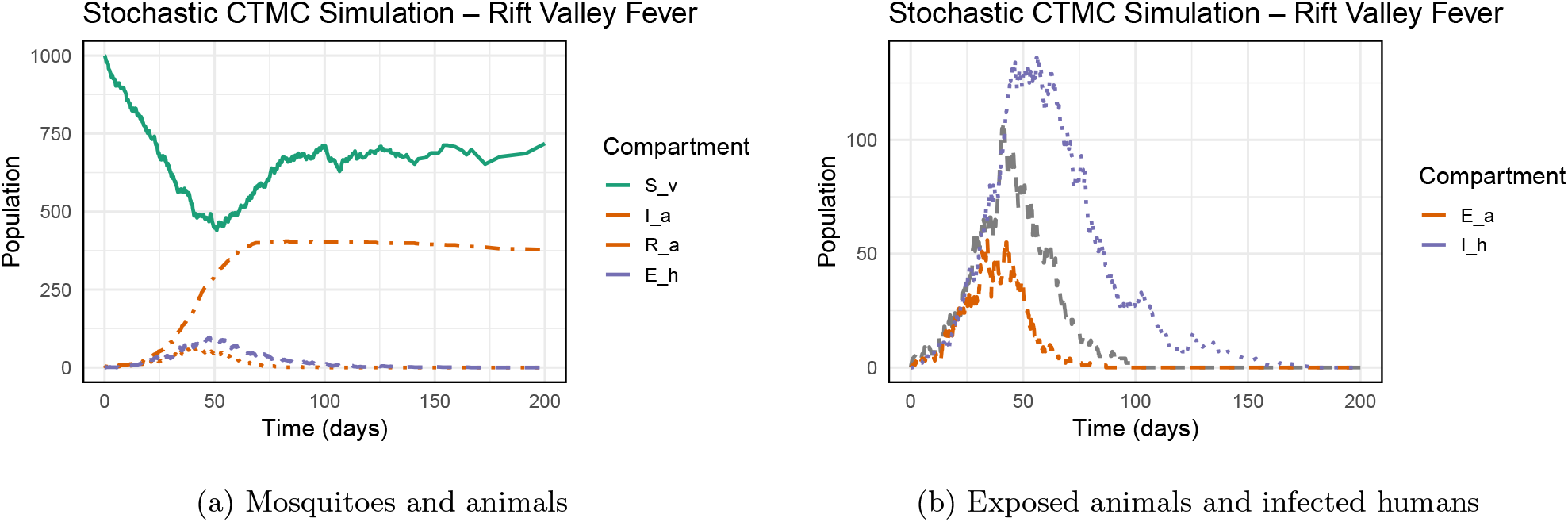
Dynamics of mosquitoes, animals, and humans in the CTMC simulation.

**Figure 5:**
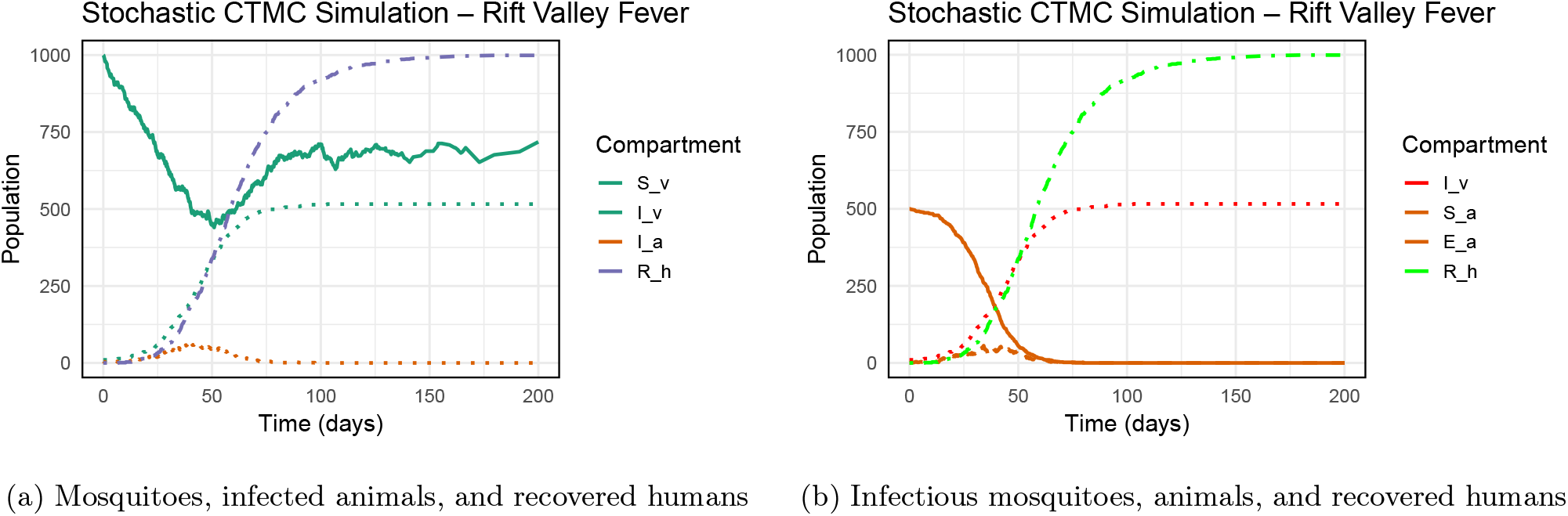
CTMC simulation showing the progression of Rift Valley Fever among vectors, animals, and humans.

In animals, the number of infectious individuals gradually increases as infectious mosquitoes transmit the virus, reaching a maximum that marks the critical point of the animal epidemic (Fig. 4a). This rise is followed by a decline due to natural recovery and disease-related mortality. The curve’s evolution helps identify optimal periods for implementing health measures, such as vaccination or herd quarantine.

Finally, the infected human population remains relatively low, and the number of recovered individuals grows slowly (Fig. 5b). This reflects the secondary role of humans in Rift Valley Fever transmission, with animal and vector dynamics remaining the main driving factors (Fig. 4b). These results (Fig. 6) highlight the importance of vector surveillance and control, as well as targeted vaccination of domestic animals in Chad, to limit disease spread and protect rural populations.

**Figure 6:**
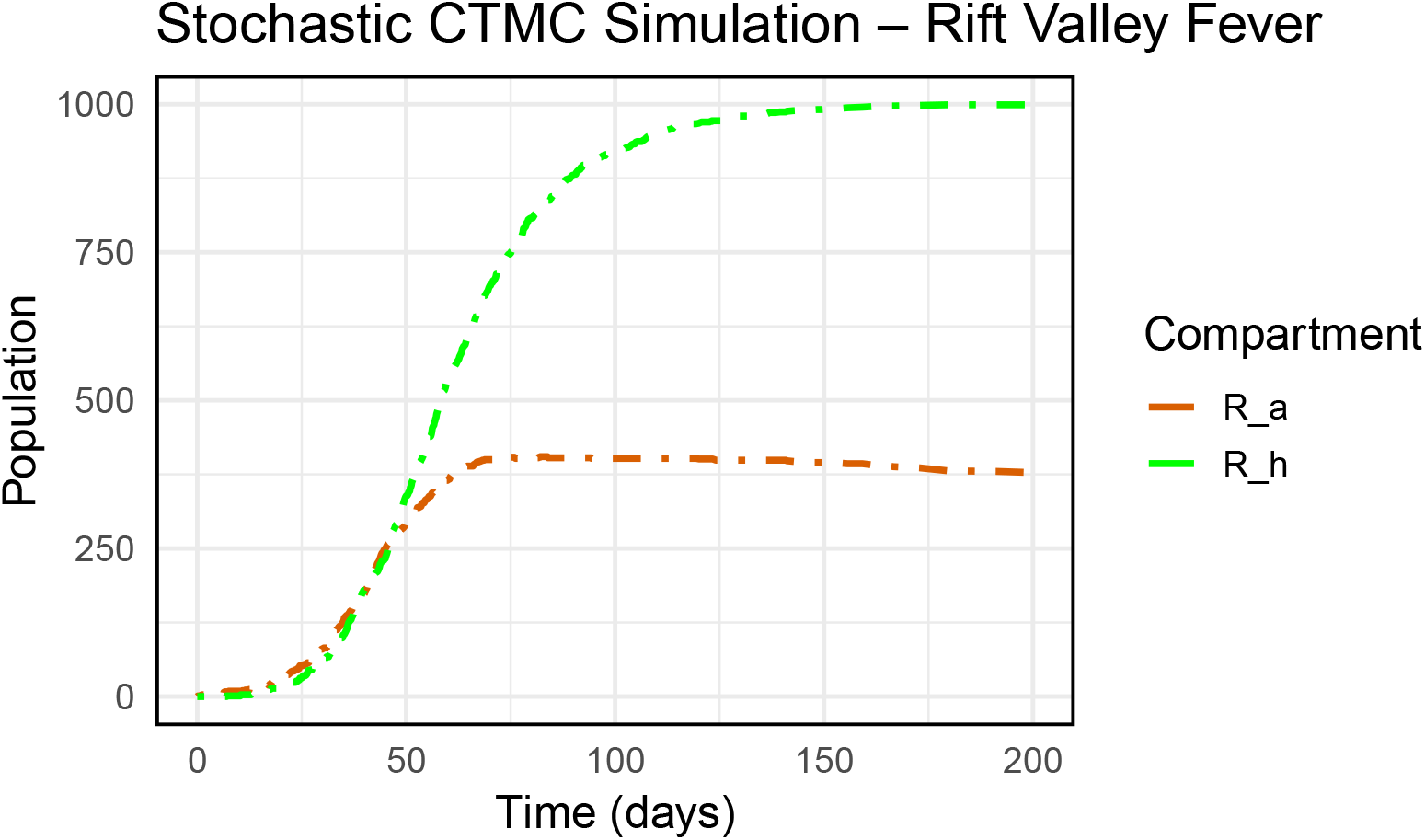
Recovered individuals (animals and humans).

### 5.4 Deterministic Simulations

Deterministic simulations are shown in Figures 7, 8, and 9. These simulations show that the populations of exposed mosquitoes, animals, and humans (Fig. 7a) and infectious individuals (Fig. 7b) gradually increase, reaching a peak around the ninth month, corresponding to the high-rainfall season in the Sahel and Chad. This season favors vector proliferation and amplifies Rift Valley Fever transmission. Exposed mosquitoes (*E*_v_) increase first, followed by infectious mosquitoes (*I*_v_), reflecting the extrinsic incubation period. Among domestic animals, exposed individuals (*E*_a_) progress quickly, followed by infectious animals (*I*_a_), indicating the critical stage of the animal epidemic. The same pattern is observed in humans.

**Figure 7:**
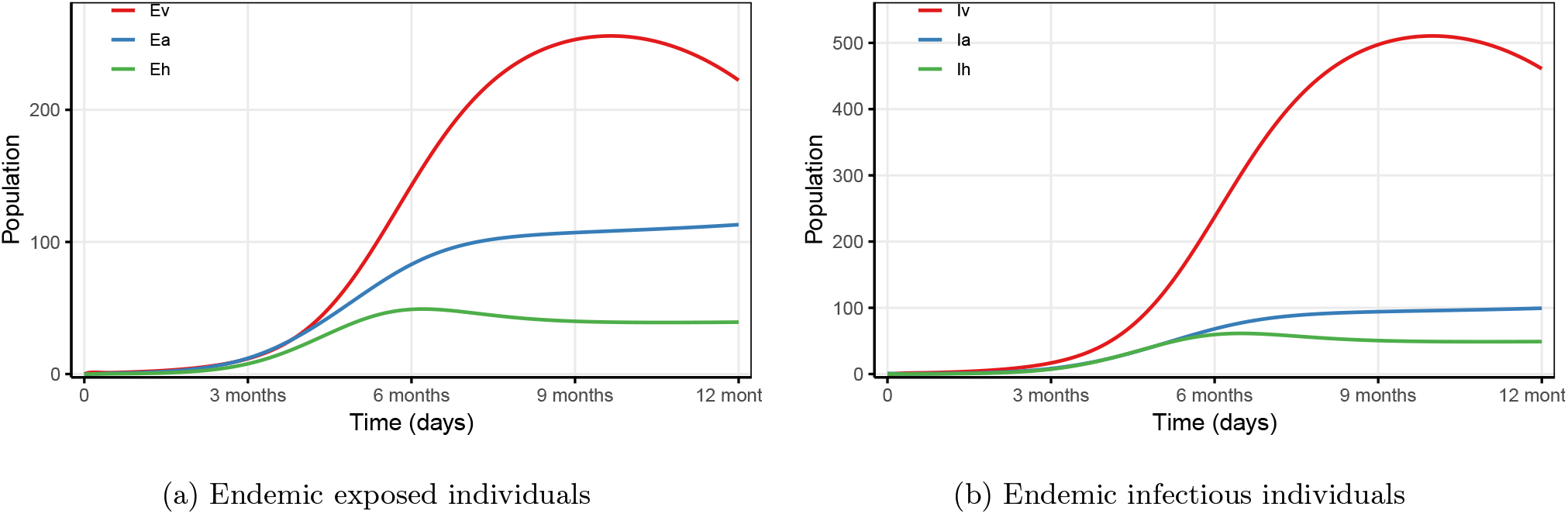
Case *R*_0_ *>* 1. The evolution of infectious individuals (b) closely follows the exposed population (a).

**Figure 8:**
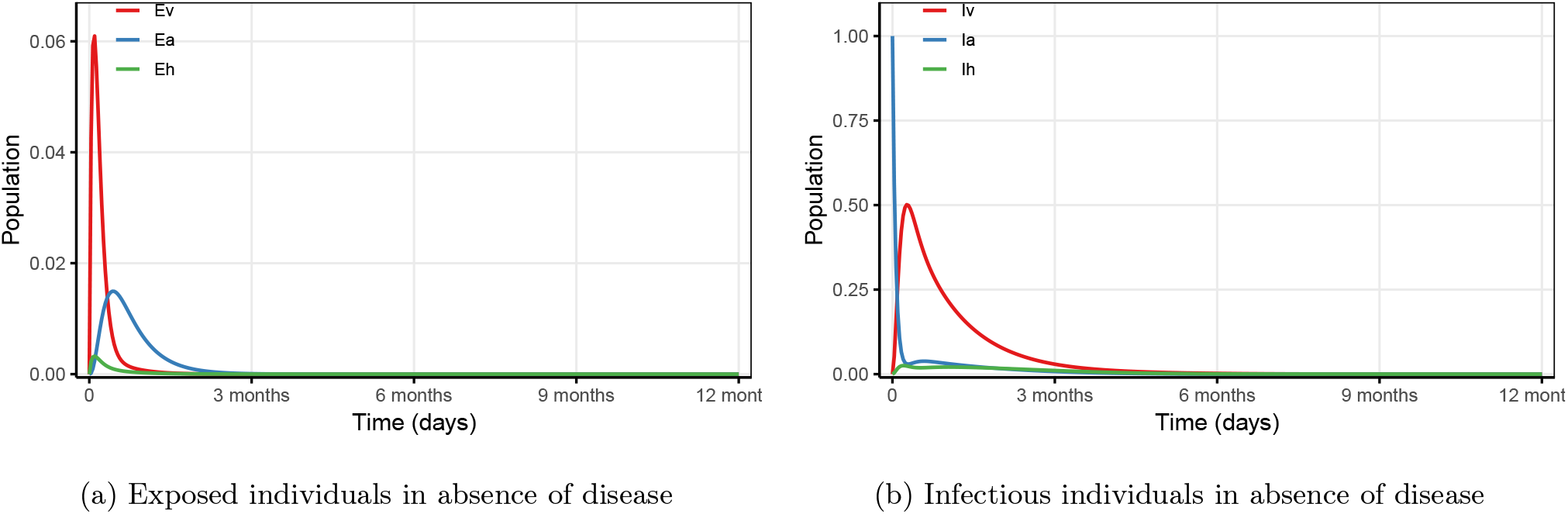
Case *R*_0_ *<* 1. Treatment of exposed (a) and infectious (b) individuals greatly reduces disease spread.

**Figure 9:**
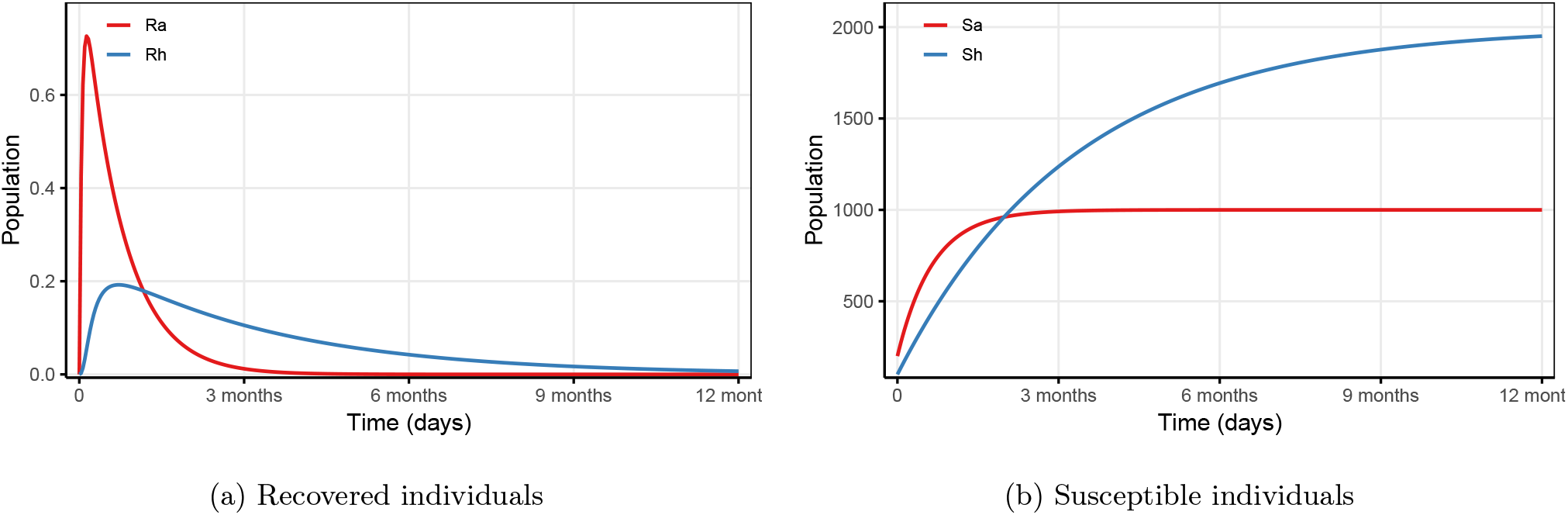
Case R_0_ *<* 1. Recovered individuals (a) result from treatment of exposed and infectious populations (see Fig. 8a and Fig. 8b). As recovered individuals become susceptible again, the susceptible population evolves accordingly (b).

The subsequent decline (Figs. 8a and 8b) is due to natural recovery and disease-related mortality (Figs. 9a and 9b).

These results highlight the key role of climatic seasonality in disease dynamics and the importance of targeting vector control and animal vaccination during the rainy season.

### Study Limitations

This study has several limitations. First, seroprevalence data [3] are cross-sectional and may not capture spatial heterogeneity. Second, the model assumes homogeneous mixing, ignoring transhumance movements and spatial structure. Third, rainfall is the only climatic factor considered; temperature and humidity are omitted. Fourth, some fixed parameters come from East African studies [8, 22] and may not fully reflect Chadian conditions. Fifth, the deterministic model cannot capture stochastic extinction events. Future work will integrate GPS tracking of livestock and spatially-explicit metapopulation dynamics.

## Conclusion

We developed a mathematical model of Rift Valley Fever integrating vector mosquitoes, ruminants, and humans, based on a *SEIR* structure with vertical transmission in vectors. Using local data from Chad (Sarh and N’Djamena, see Fig. 2) allowed us to illustrate the differential impact of Sudanian and Sahelian climates on mosquito dynamics and disease spread.

The mathematical analysis (Section 3) demonstrated the positivity of the model, enabled the determination of the basic reproduction number R_0_, and established the local and global stability of the disease-free equilibrium. The calibrated model yielded R_0_ = 2.16 (95% CI: 1.89–2.43), confirming active transmission in the Sahelian zone. The sensitivity analysis (Section 5.2) identified the most influential parameters for transmission dynamics.

Deterministic simulations (Section 5.4) and the stochastic continuous-time Markov chain approach (Section 5) confirmed the critical role of seasonal precipitation, with peaks of animal and human infections around the ninth month, corresponding to heavy rainfall periods.

This work highlights the importance of an integrated “One Health” approach for surveillance, prevention, and planning of interventions against Rift Valley Fever in Chad.

## Data Availability

The rainfall data used in this study were obtained from the National Meteorology Agency of Chad (ANAM-TCHAD). Seroprevalence data were obtained from previously published studies cited in the manuscript. All other data generated during this study are included in the manuscript. The R code used for numerical simulations is available from the corresponding author upon reasonable request.

